# Miniaturized microscope for non-invasive imaging of leukocyte-endothelial interactions in human microcirculation

**DOI:** 10.1101/2023.07.21.23292515

**Authors:** Arutyun Bagramyan, Charles P. Lin

**Affiliations:** Wellman Center for Photomedicine and Center for Systems Biology, Massachusetts General Hospital, Harvard Medical School, Boston, MA, USA

## Abstract

*We present a miniature oblique back-illumination microscope (mOBM) for imaging the microcirculation of human oral mucosa, enabling real-time, label-free phase contrast imaging of leukocyte rolling and adhesion, the initial steps in leukocyte recruitment that is a hallmark of inflammation. Imaging cell motion can provide new diagnostic information (time course of disease progression, response to therapy, etc*.*) that is not available using traditional static diagnostic parameters such as cell number and morphology*.

## MAIN TEXT

The ability of leukocytes to traffic to various organs and tissues is a fundamental requirement for proper immune function^1^. Leukocyte recruitment is a multi-step process initiated by the slowdown of circulating leukocytes that tether and roll on the endothelial surface, followed by their firm adhesion and extravasation into tissue^2^. Though well characterized in animal models using intravital microscopy, the rolling and adhesion events (collectively known as leukocyte-endothelial interaction, or LEI^3,4^) have rarely been observed in humans^5^. Conceptually, imaging cell motion as a potential source of diagnostic information has yet to be explored in clinics, as traditional histopathology has relied on the static examination of biopsied samples. LEI is reported to be significantly increased in the sublingual microvasculature of patients with systemic inflammation such as sepsis^6^ and ischemia-reperfusion injury^7^. However, assessing LEI in clinical settings has been challenging due to the lack of proper detection and analytical tools. Individual leukocytes are not resolved using existing clinical instruments such as CytoCam and MicroScan^8^; instead, their presence is inferred from the gaps or voids in the blood vessels otherwise filled with red blood cells. The suboptimal image quality is further compounded by the severe motion and pressure artifacts and a lack of proper analytical tools to quantify leukocyte motion^9^). Reflectance confocal microscopy (RCM) provides high-resolution cellular imaging and has been successfully used in dermatology clinics^10^. However, RCM requires laser scanning with limited frame rate^11^. Nonlinear optical techniques such as third-harmonic generation (THG) microscopy^12^ and two-photon-induced UV autofluorescence imaging^13^ can also provide label-free imaging of leukocytes but these techniques entail complex laser systems and scanning platforms with no clear path for translation to the bedside.

To address these limitations, we developed a miniaturized oblique-back illumination microscope (mOBM) for non-invasive imaging of the microvasculature of human oral mucosa (Fig. 1a-c). The mOBM (Fig. 1d) consists of a 1 mm diameter aberration-corrected gradient index (GRIN) objective lens with a numerical aperture (NA) of 0.75 and a green LED (light emitting diode) light source coupled to a large core (Ø 1mm) multimode optical fiber. The output end of the fiber is positioned such that photons enter the tissue from one side of the GRIN lens (Fig. 1e). Injected photons undergo multiple scattering events in deep tissue layers, and only a fraction of the photons are collected by the GRIN lens, providing the back-illumination at an oblique angle due to the offset geometry of the optical fiber^14^. The asymmetric (oblique) back illumination generates phase-gradient contrast (PGC) images that resemble differential interference contrast (DIC)^15^ or differential phase contrast (DPC)^16^ microscopy images with a crucial difference. Unlike DIC or DPC, OBM is designed to work with intact, thick tissue^14^ and is therefore compatible with *in vivo* imaging. The use of the green LED additionally provides contrast based on hemoglobin absorption, making the white blood cells stand out against the darker red blood cells (Fig. 2a, left).

**Fig. 1.**
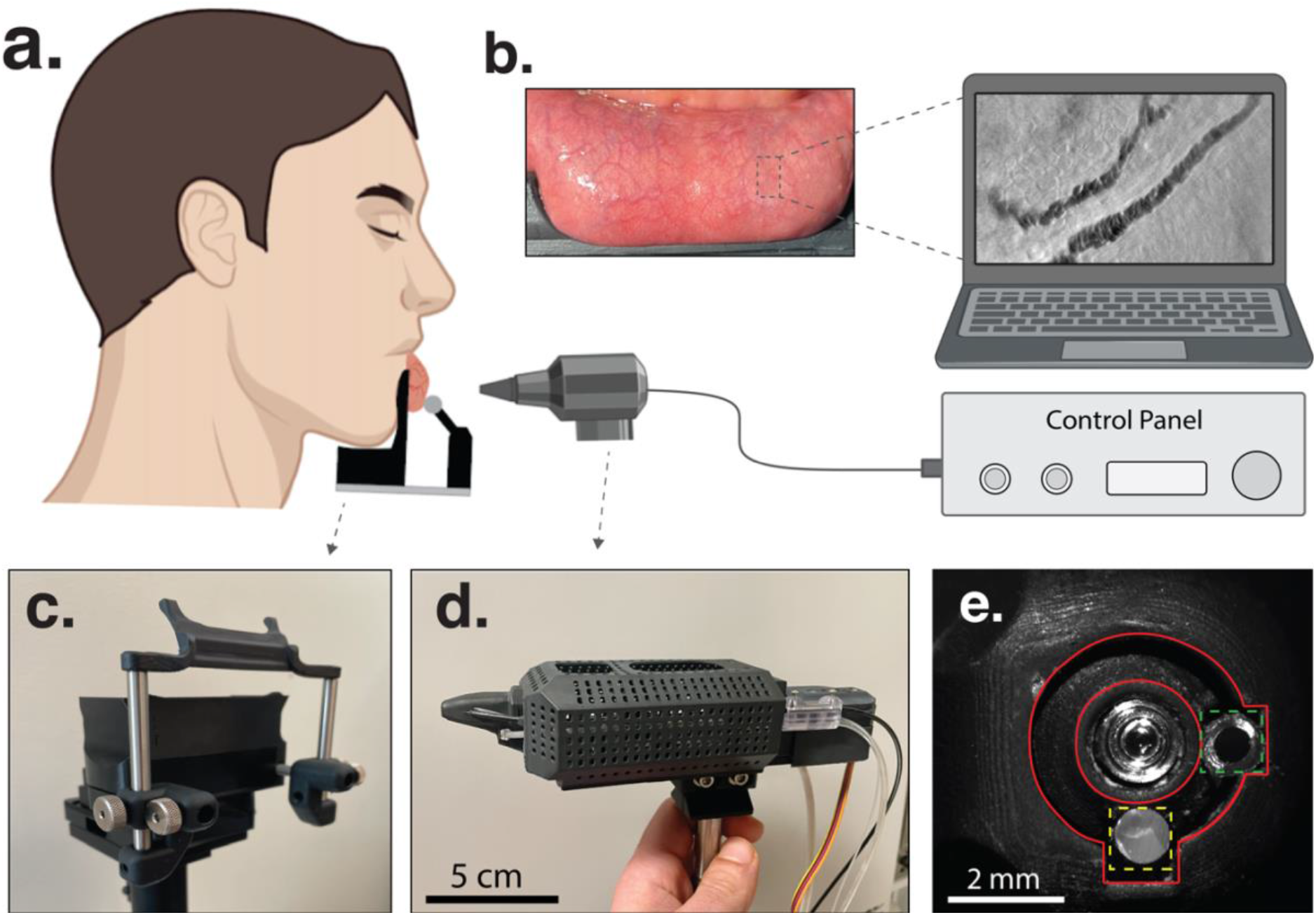
System for *in vivo* imaging of human oral mucosa. **a**. Schematics of the human set-up. **b**. Exposed oral mucosa tissue of a healthy human subject. The microvasculature is shallow and easy to access with our imaging instrument presented in (d). **c**. Oral mucosa apparatus. **d**. The developed mOBM. **e**. mOBM’s imaging tip: GRIN lens (center), optical fiber (yellow dotted square), immersion medium tube (green dotted square), and the vacuum cavity (area between red circles).

**Fig. 2.**
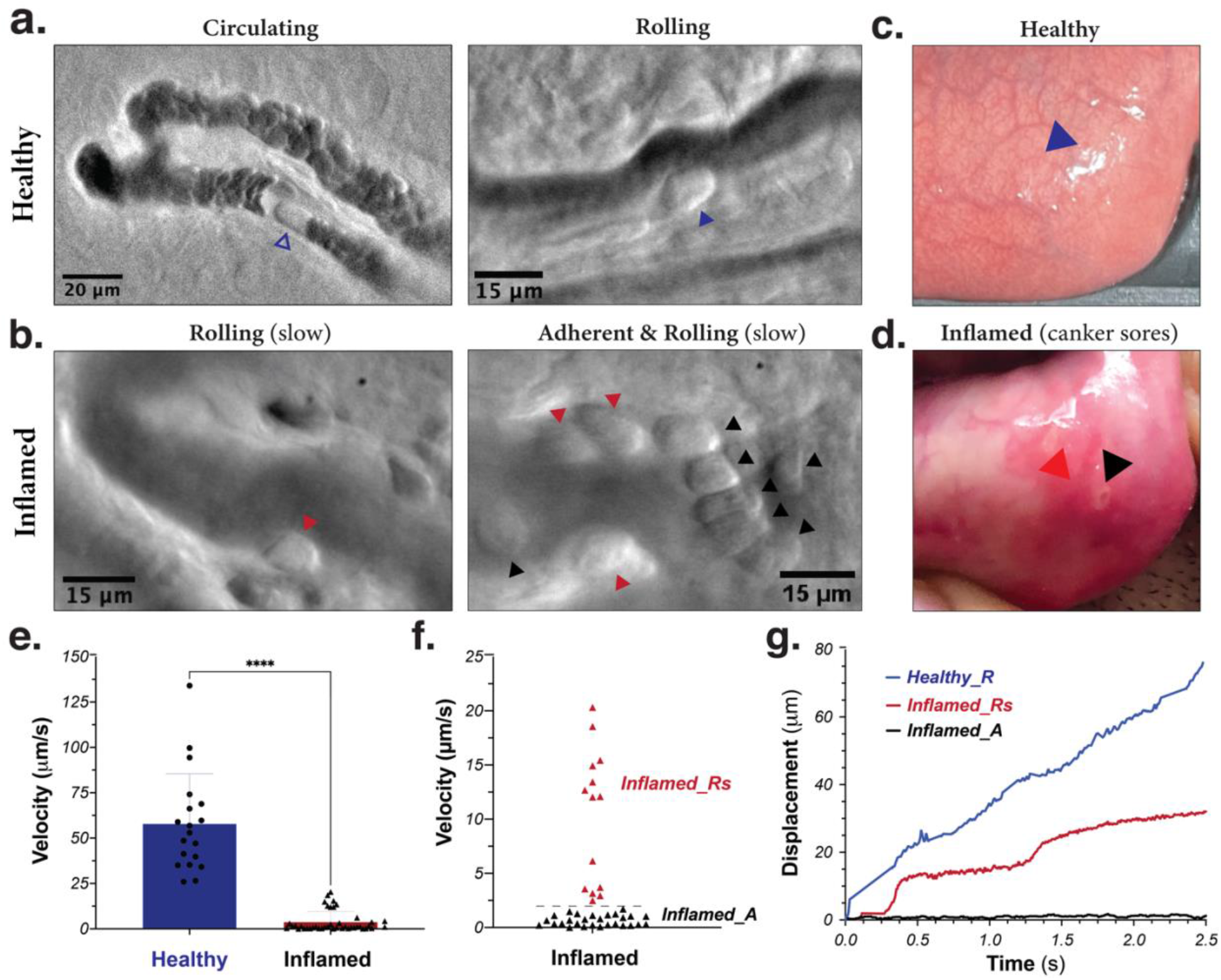
LEIs in human microvasculature. a. Healthy condition. *Left:* A circulating leukocyte (open blue arrowhead); single frame from Supplementary Video 1. *Right:* A rolling leukocyte (blue arrowhead); averaged image of five single (consecutive) frames from Supplementary Video 2. **b. Inflamed condition**. *Left:* A slow-rolling leukocyte (red arrowhead). *Right:* Slow-rolling and adherent leukocytes (red and black arrowheads, respectively) from Supplementary Video 3. **c-d**. Healthy and inflamed oral mucosa tissue. **e-g**. Velocity and displacement profiles of individual leukocytes. In ‘‘e’’, we have used unpaired t-test (blue column, n=18; Red column, n=39). P-value<0.0001. \***Note:** Images of the inflamed tissue were obtained with an earlier version of the instrument (Supplementary Fig. 1). Data acquisition parameters and image processing pipelines are detailed in *Online Methods*.

To minimize motion and pressure artifacts, a custom tissue stabilizer (Fig. 1c) is developed to gently expose the lower lip’s inner microvasculature (Fig. 1b) and to maintain its position for the duration of the imaging. The choice of the inner lip is based on its ready access, its well-developed shallow vascular bed, its lack of skin pigmentation, the absence of the highly scattering stratum corneum layer that degrades the image quality in the skin, and the comfort of participants. To validate the performance of our system we imaged the movements of leukocytes in the mucosal microvasculature of human volunteers. When imaged at a frame rate of 200 Hz, individual blood cells in the microcirculation are clearly delineated (Fig. 2a, left; Supplementary Video 1). These cells travel at a speed of ∼1 mm/sec, and their motions are effectively “frozen” when the exposure time of individual frames is set to <1 msec. In addition, rolling cells are detected in a subset of blood vessels of healthy volunteers (Fig. 2a, right; Supplementary Video 2), likely postcapillary venules based on their diameter and flow speed.

We also imaged an inflamed area caused by the presence of canker sores (Fig. 2b-d; Supplementary Video 3), and observed a drastic reduction in the average speed of leukocytes. Using automated frame-by-frame leukocyte tracking (Online Methods), we obtained an average rolling velocity of 58 ± 28 µm/s in the healthy tissue (Fig. 2e), which reduced to 4 ± 6 µm/s in the inflamed tissue (Fig. 2e). Closer inspection revealed two populations (Fig. 2f), an adherent population with an average velocity ≈ 0.7 ± 0.7 µm and a slowly rolling population with an average velocity of 11 ± 6 µm. The adherent cells were restricted to the area close to the canker sores (Fig. 2d, black arrowheads), while the slowly moving leukocytes were detected both in the center and in the periphery (Fig. 2d, red arrowheads) of the inflamed area. The displacement profile showed the characteristic stop-and-go movement patterns during inflammation (Fig. 2g).

To make the instrument even more compact, the current focusing mechanism (a translational stage) can be replaced with an electrically tunable lens^17^. In addition, the high-speed CMOS sensor (frame rate up to 1000 Hz) can be replaced by a standard (30 Hz) video camera that will still be able to image rolling and adherent cells, but the flowing cells will not be resolved at this frame rate. The high-speed imaging capability offers the tantalizing possibility of performing noninvasive white blood cell count by resolving individual circulating cells and flagging leukocytes “on the fly” with the help of machine learning, a subject of active pursuit in our laboratory and others^18^. Because of its compact size, low cost, and simple construction, we expect to place the mOBM instrument in several clinics to begin testing the diagnostic utility in critically ill patients and preterm infants at high risk of infection and sepsis. We further envision the instrument to find use in resource-poor settings that lack the expertise and infrastructure to draw blood for standard laboratory analysis.

## Data availability

The data that support the findings of this study are available from the corresponding author upon reasonable request.

## Ethics statement

The experimental protocols related to human volunteers were performed in accordance with the guidelines and regulations of Massachusetts General Hospital. The study protocol (#2021P003047) was approved by the Internal Review Board (IRB) of Massachusetts General Hospital. Informed consent was obtained from all subjects participating in our study.

## REFERENCES

1. Germain, R. N., Robey, E. A. & Cahalan, M. D. A Decade of Imaging Cellular Motility and Interaction Dynamics in the Immune System. Science 336, 1676–1681 (2012).

2. Vestweber, D. How leukocytes cross the vascular endothelium. Nat. Rev. Immunol. 15, 692–704 (2015).

3. Springer, T. A. Traffic signals for lymphocyte recirculation and leukocyte emigration: The multistep paradigm. Cell 76, 301–314 (1994).

4. von Andrian, U. H. et al. Two-step model of leukocyte-endothelial cell interaction in inflammation: distinct roles for LECAM-1 and the leukocyte beta 2 integrins in vivo. Proc. Natl. Acad. Sci. 88, 7538–7542 (1991).

5. Sahu, A. et al. In vivo tumor immune microenvironment phenotypes correlate with inflammation and vasculature to predict immunotherapy response. Nat. Commun. 13, 5312 (2022).

6. Fabian-Jessing, B. K. et al. In vivo quantification of rolling and adhered leukocytes in human sepsis. Crit. Care 22, 240 (2018).

7. Uz, Z., Ince, C., Shen, L., Ergin, B. & van Gulik, T. M. Real-time observation of microcirculatory leukocytes in patients undergoing major liver resection. Sci. Rep. 11, 4563 (2021).

8. Massey, M. J. & Shapiro, N. I. A guide to human in vivo microcirculatory flow image analysis. Crit. Care 20, 35 (2016).

9. Ince, C. et al. Second consensus on the assessment of sublingual microcirculation in critically ill patients: results from a task force of the European Society of Intensive Care Medicine. Intensive Care Med. 44, 281–299 (2018).

10. Ulrich, M., Lange-Asschenfeldt, S. & Gonzalez, S. The use of reflectance confocal microscopy for monitoring response to therapy of skin malignancies. Dermatol. Pract. Concept. 2, 0202a10 (2012).

11. Rajadhyaksha, M., González, S., Zavislan, J. M., Rox Anderson, R. & Webb, R. H. In Vivo Confocal Scanning Laser Microscopy of Human Skin II: Advances in Instrumentation and Comparison With Histology11The authors have declared conflict of interest. J. Invest. Dermatol. 113, 293–303 (1999).

12. Wu, C.-H. et al. Imaging Cytometry of Human Leukocytes with Third Harmonic Generation Microscopy. Sci. Rep. 6, 37210 (2016).

13. Li, C. et al. Imaging leukocyte trafficking in vivo with two-photon-excited endogenous tryptophan fluorescence. Opt. Express 18, 988–999 (2010).

14. Ford, T. N., Chu, K. K. & Mertz, J. Phase-gradient microscopy in thick tissue with oblique back-illumination. Nat. Methods 9, 1195–1197 (2012).

15. Nomarski G. Differential microinterferometer with polarized waves. J Phys Radium Paris 16, 9S (1955).

16. Hamilton, D. K. & Sheppard, C. J. R. Differential phase contrast in scanning optical microscopy. J. Microsc. 133, 27–39 (1984).

17. Bagramyan, A. et al. Focus-tunable microscope for imaging small neuronal processes in freely moving animals. Photonics Res. 9, 1300–1309 (2021).

18. Huang, L., McKay, G. N. & Durr, N. J. A Deep Learning Bidirectional Temporal Tracking Algorithm for Automated Blood Cell Counting from Non-invasive Capillaroscopy Videos. in Medical Image Computing and Computer Assisted Intervention – MICCAI 2021 (eds. de Bruijne, M.et al.) 415–424 (Springer International Publishing, 2021). doi:10.1007/978-3-030-87237-3_40.

