## Supplementary Materials for "Miniaturized microscope for non-invasive imaging of leukocyte-endothelial interactions in human microcirculation"

### Online Methods*:*

**Key Features of the Developed System.** The optical design of mOBM was accomplished using the Zemax software (Supplementary Figure 2). It consists of a 1 mm diameter gradient index (GRIN) objective lens, relay optics, and a CMOS camera. The imaging GRIN lens assembly (Grintech, *GT-MO-080-032-ACR-VISNIR-08-20*) is aberration-corrected and has a high numerical aperture (NA) of ≈0.75 (in water). The relay optics consists of a GRIN lens (Edmund Optics, *#64-519*) with a ≈1/4 pitch and NA=0.52 together with an achromat doublet lens (Edmund Optics, *#49-772*) with an effective focal length of 19.1 mm that projects the magnified image onto the CMOS sensor (Basler, *daA1920-160um*). Oblique illumination is provided by a 530 nm LED light source (Thorlabs, *M530F2*) coupled to a 1 mm diameter large core optical fiber, whose output (≈ 19 mW) end is positioned on one side of the GRIN objective lens assembly (Fig. 1e). The developed mOBM (Fig. 1d) detects multiply scattered photons, in a non-confocal manner, generating phase-gradient contrast (PGC) images that reveal fine morphological details (e.g., cell membrane, granules). In addition to the PGC, our instrument also benefits from absorption-contrast generated by using the 530 nm wavelength that is strongly absorbed by hemoglobin (in red blood cells) and not by leukocytes. An important characteristic is the imaging tip (Fig. 1e), which consisted of four channels: **1.** A central channel containing the miniaturized GRIN objective lens. **2.** An illumination channel containing the large core multi-mode fiber. **3.** An irrigation channel to keep oral mucosa tissue moist and to maintain the immersion medium in front of the objective lens. **4.** A ring-shaped vacuum cavity surrounds the central channel to stabilize the imaging area. The magnification of our instrument was ≈ 25, which resulted in ≈ 7.2 pixels/µm (without binning) and ≈ 3.6 pixels/µm (with 2x2 binning) ratios. The field-of-view of our system was ≈ 150x200 um and the working distance of the objective lens was adjusted (20-100 µm) by means of motorized displacement of the camera in the direction of the optical axis. SolidWorks software was used to design mOBM's mechanical housing, the imaging tip, and the oral mucosa apparatus (Fig. 1c). Parts were printed using a 3D laser printer (Formlabs 3B). All parts in contact with the human tissue were printed with sterilizable, biocompatible materials (Formlabs, *RS-F2-BMCL-01, RS-F2-BMBL-01*).

**Tissue Stabilizer.** The developed universal oral mucosa apparatus (Fig. 1c) was used to gently hold the subject's lower lip tissue and expose buccal microvasculature (Fig. 1b) without generating undesired blood flow alternations. The apparatus also stabilized the tissue for the duration of the imaging session (≈20-60 min). A motorized XYZ stage was used to position the imaging tip of the mOBM and locate the microvasculature of interest.

**Imaging of Healthy Participant.** The experimental protocols related to human volunteers were performed in accordance with the guidelines and regulations of Massachusetts General Hospital. The study protocol (#2021P003047) was approved by the Internal Review Board (IRB) of Massachusetts General Hospital. The imaging session and data collection started after the subjects had signed the IRB-approved consent form. Subjects were first seated in front of the imaging system and their head was gently stabilized using soft tissue straps. The lower lip was then enrolled using the developed oral mucosa apparatus (Fig. 1c). There were no additional requirements from the subjects other than remaining in a seating position for the duration of the imaging. The imaging session was initiated by the operator who positioned the imaging tip over the region of interest (ROI) using a motorized XYZ actuator with micrometer resolution. The imaging session usually lasts between 20-60 minutes.

**Image acquisition parameters.** For the circulating leukocytes (Fig. 2a, left), the acquisition frame rate was fixed at 200 fps and the exposure time varied between 0.5-1 ms. If necessary, 2x2 binning was used to increase the signal-noise ratio. For the rolling leukocytes (Fig. 2a (middle) and Fig. 2b), the acquisition frame rate was fixed at 200 fps and the exposure time varied between 2.5 - 4 ms.

**Image Processing Pipeline.** Image processing was performed in *ImageJ* (open-source software). The image processing pipeline for the rolling and adherent leukocytes: 1. Registration (plug-in: *Template Matching).* 2. Cropping the region of interest (ROI). 3. Blood flow smoothing (plug-in: *Kalman filter*). 4. Extraction of PGC (Image - Gaussian blurred image (sigma radius: 20-40)). 5. Average subtraction (PGC stack – averaged image) 6. Leukocyte tracking (plug-in: *TrackMate*). Leukocytes that detached from the endothelial wall after brief contact(s) were excluded from the analysis. The image processing pipeline for the circulating leukocytes: 1. Registration (plug-in: *Template Matching).* 2. Cropping the ROI. 3. Extraction of PGC (Image - Gaussian blurred image (sigma radius: 20-40)).

**Statistical analysis.** For the statistical analysis (Fig. 2e) we have used the unpaired t-test method (also known as the student t-test). In the rolling cell group (Fig. 2e, blue column), n=18; Inflamed group (Fig. 2e, red column), n=39. P-value <0.0001.

### Supplementary Videos:

The videos can be downloaded at:
https://www.dropbox.com/sh/jkcdtwmu53wzf8q/AABqV7i_iAeWFE4uQ8UOz74xa?dl=0

**Supplementary Video 1:** Circulating leukocytes in the oral mucosa microvasculature of a healthy volunteer. Acquisition frame rate: 200fps.

**Supplementary Video 2:** Rolling leukocytes in healthy oral mucosa tissue. Acquisition frame rate: 200fps.

**Supplementary Video 3:** Adherent and slowly rolling leukocytes in inflamed oral tissue. Acquisition frame rate: 200fps.

### Supplementary Figures:

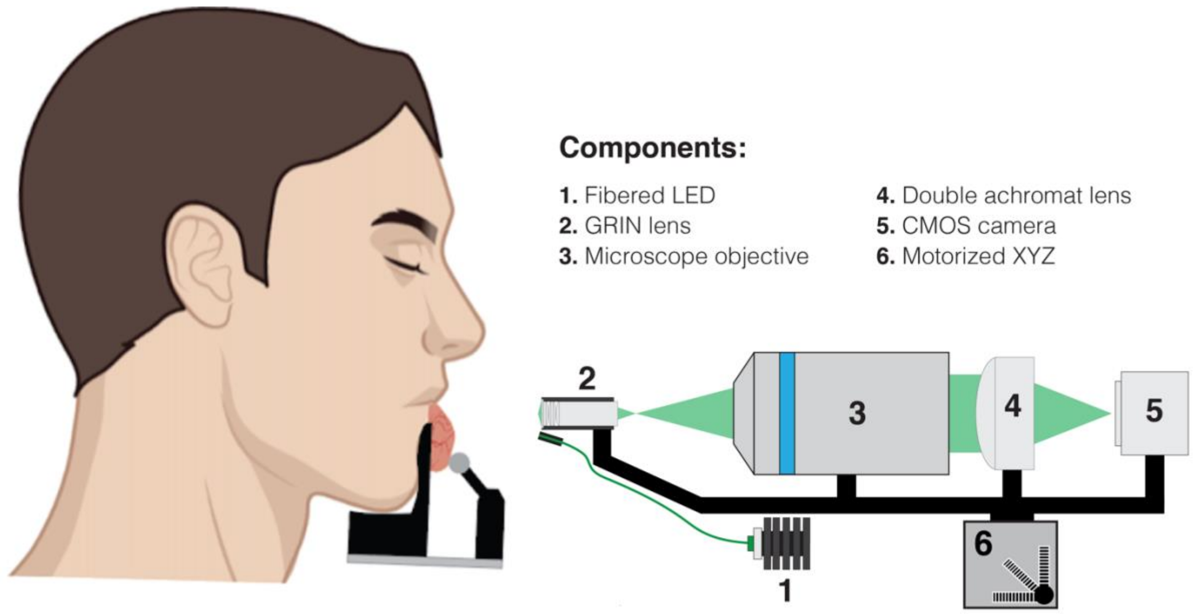


**Supplementary Figure 1 |** An earlier version of the mOBM system for oral mucosa imaging in human subjects that uses a microscope objective (Mitutoyo, *378-804-3*) was later replaced with a GRIN lens (Supplementary Figure 2, component #2).

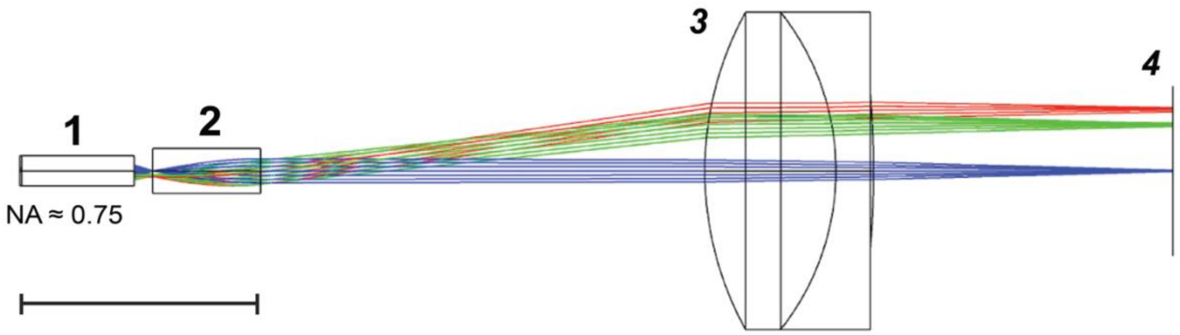


**Supplementary Figure 2 |** Optical simulation (Zemax) of the imaging pathway of the miniaturized instrument (Fig. 1d). Components: 1. Miniaturized aberration-corrected GRIN lens assembly, NA ≈ 0.75 (in water); 2. Standard GRIN lens, N=0.52; 3. Achromat doublet lens, EFL = 19.1 mm 4. Monochrome CMOS camera.
